# Patient engagement in research; benefits, challenges, importance, and implications

**DOI:** 10.1101/2023.03.28.23287870

**Authors:** Caitlyn Ivany, Tess Hudson, Patricia Schneider, Hadia Farrukh, Marilyn Swinton, Michelle Ghert

## Abstract

**Objective:** Patient engagement (PE) and patient-oriented research have begun to reshape the thought process behind conducting research with the aim of maximizing the relevance of findings for patients. This study aimed to examine the perceived benefits, challenges, importance, and implications of patient engagement from the perspectives of sarcoma patient advisors and researchers.

**Methods:** This study utilized a mixed model design. Qualitative data was collected through two focus group discussions with sarcoma patients. Quantitative data was collected via a survey containing Likert scale questions completed by the Centre for Evidence-Based Orthopaedics Musculoskeletal Oncology research team at McMaster University.

**Results:** Results showed that patients value the opportunity to contribute to research and support future patients. Being a patient advisor also creates a sense of community and fosters support through building connections and communicating with other patients. Members of the research team noted that patient engagement is important for the study of patient relevant topics and provides insight into the improvement of patient care. However, an added challenge is the lack of current guidance surrounding the implementation of patient engagement.

**Conclusion:** These findings emphasize the potential value of patient engagement while also highlighting the need for further research into best practices for the implementation of patient engagement efforts. Overall, patient engagement is an essential area in need of further exploration to enhance future research and clinical trials.

## 1.0 Introduction

The involvement of patients in research has become a widely discussed topic in the field of healthcare. As noted in prior literature, it is common for patients and researchers or healthcare practitioners to have different views of study priorities pertaining to a certain health condition, disease, treatment, etc. [1]. Involving patients in research that pertains to their health provides the opportunity to incorporate a patient perspective and comes with many benefits including improved health outcomes, patient empowerment, and increased relevance of research findings [1-4]. However, there is still work to be done to further establish the importance and value of patient engagement. This current study aims to explore patient engagement from multiple perspectives with the objective of outlining the benefits, challenges, importance, and implications of patient engagement efforts in research.

### 1.1 Defining Patient Engagement

‘Patient engagement’ is best explained by examining each word individually. In the context of patient engagement, the term ‘patient’ generally refers to anyone with personal experience with the health condition, disease, or treatment that the research focuses on. This demographic can therefore include patients, caregivers, and family members [1, 5]. ‘Engagement’ refers to active involvement in research [1, 5]. ‘Patient engagement’ therefore refers to the active participation of patients in research and in collaboration with a research team or healthcare provider.

Engagement encompasses an array of tasks that involves patients in various aspects of the research process such as developing the study topic, research question, methodology, and translation of results [1, 6]. Higgins and colleagues analyzed scientific literature to define four main attributes of patient engagement: personalization, access, commitment, and therapeutic alliance [7]. Personalization refers to the need for engagement to be tailored to a patient’s specific needs for research findings to be valuable [7]. Access refers to the patient’s ability to understand the necessary information to successfully contribute and provide their experience to the study [7]. Commitment entails the patient’s drive to be part of the research process and their willingness to collaborate [7]. Lastly, therapeutic alliance describes the relationship built between the patient and healthcare provider throughout the engagement process with the common goal of improving health outcomes [7]. These four attributes outline the basic components of patient engagement and provide a foundation of understanding for the definition of patient engagement.

### 1.2 Current Patient Engagement Strategies

Although the most effective methods of patient engagement have yet to be explicitly defined, some researchers have outlined broad guidelines on developing a successful research partnership with patients. Kirwan and colleagues proposed that these guidelines include the development of supportive policies to maintain organization, fostering positive attitudes and respect between patients and researchers, meeting training requirements to ensure baseline knowledge of research is acquired, and identifying areas that require advanced planning to ensure success [8]. Hewlett and colleagues initially proposed similar guidelines in their development of the acronym FIRST (Facilitate, Identify, Respect, Support, Train) to guide patient engagement [2]. Aside from these common principles, a prevalent component of patient engagement is education. Multiple studies have demonstrated the importance of education and engagement in the form of frequent doctor visits as follow-up and providing information regarding the signs of disease to allow for early detection of disease recurrence [9, 10]. As well, providing background knowledge is an important aspect of engaging patients as research partners. Knowledge of the scientific process and the study topic provides patients with the tools to feel that they are able to contribute valuable information and actively participate in research decisions [8].

To select patients for engagement, many studies report using convenience sampling and creating advertisements to recruit volunteer patients [11]. Random sampling would be the ideal method of selecting patients to eliminates bias. However, random sampling is often difficult with patient engagement research due to the low number of chosen patients and, depending on the study topic, the health condition being researched [11]. Researchers commonly use focus groups, interviews, and surveys throughout the research process to engage patients and hear their perspective [11].

Various organizations have developed initiatives to increase the prevalence of studies involving patients such as the Canadian Institutes of Health Research (CIHR) Strategy for Patient-Oriented Research (SPOR) program initiated in 2014 [12]. The objective of this program is to engage patients to ensure research focuses on relevant priorities from the patient perspective [1]. With the guidance of this framework, the CIHR hoped to facilitate collaboration between patients, researchers, and healthcare decision makers to improve Canadian healthcare policies and practices in sustainability, accessibility, and equity [6]. To accomplish the program’s objectives, centers of expertise have been developed at the provincial, territorial, and national level with allocated research funds for patient-oriented research efforts [6]. The SPOR Capacity Development Framework provides SPOR partners and researchers with a cohesive vision and fundamental principles of patient engagement [6]. The framework emphasizes training, mentoring, and career support to guide researchers through engagement efforts [6]. Examples of engagement under this framework include providing patients with the necessary tools to contribute to research, including patients in administrative roles and decision making, planning events for patients, and consistently highlighting the value of patient perspective in research [6]. In addition, the CIHR posits inclusiveness, support, and mutual respect as their guiding principles of their SPOR program [6]. A Canadian study under the direction of the SPOR initiative implemented patient engagement to ensure the needs of patients were addressed in inflammatory bowel disease (IBD) research. The study recruited patient partners with lived experience to gain insight on why individuals choose to participate in research and if there are any barriers to their participation. The patient partners in this study were trained in qualitative research methods, helped to develop interview guides, and assisted with data analysis by identifying common themes in patient interview responses [6]. The study concluded that patients are most inclined to participate in research when engagement efforts are convenient, flexible in terms of scheduling, organized, involve compensation, and when researchers regularly communicate the study’s progress [6]. The development of Canada’s SPOR emphasizes the importance of patient engagement efforts and provides guiding practices for researchers looking to utilize this novel study approach. However, further research is needed to fully outline the impact of and best practices for successful patient engagement.

### 1.3 Engagement at Different Stages of the Research Process

Research has explored patient engagement at various stages of the research process to determine where it is most effective and beneficial. Many existing studies that employ patient engagement do not sustain engagement efforts throughout the duration of the study and mainly collaborate with patients only at the start. This is often due to time and money constraints [3]. Other researchers have also found that engagement is preferred during the initial preparation phase and the final dissemination and knowledge translation phase, although engagement throughout is ideal [13]. Brett and colleagues found that patient engagement had an impact on all stages of the research process [14]. In the initial stages of research, patients were helpful in identifying relevant topics based on their experience. During data collection, patients were willing to offer their opinions on the study design and helped with study translation to ensure accessibility for a lay audience [14]. During the analysis and write-up phase, patients were helpful in identifying themes in the results and ensured that these themes were valuable to them [14]. Lastly, patients were able to help with spreading knowledge and implementing findings [14]. In summary, patient engagement is feasible at all time points of a research study and has been found to be most beneficial when engagement is continuous throughout the research process.

### 1.4 Current Patient Engagement Knowledge – Benefits, Challenges, Implications, and Importance

Previous research has noted many benefits of patient engagement including improved health outcomes, patient empowerment, and increased relevance of research findings [1-4]. One of the most prominent benefits noted in the literature is the value of the patient perspective. Engaging patients allows researchers to gain a new point of view from an individual with lived experience when conducting a research study [1]. Including this patient perspective improves the applicability of research outcomes, as patients with lived experience have an enhanced viewpoint as to what topics of health research would be relevant and beneficial to them and future patients with similar health conditions [3]. Researchers also benefit from PE through increased study enrolment and decreased attrition as patients tend to feel more inclined to participate in studies if the findings are valuable to them [3]. In terms of benefits for the patient, engaging with research provides patients with knowledge and a feeling of empowerment in their healthcare decisions and needs [1, 3, 4, 15]. With PE, patients are provided with the unique opportunity to develop their own voice in the research world which can also enhance their relationship with the health care system and their healthcare professionals [3, 15].

Despite numerous benefits, implementation of PE may also come with challenges such as added cost, time, and complexity. Involving patients as research partners requires training so patients can better understand the research process and be able to contribute their perspective effectively. Training is also needed for researchers to learn strategies to effectively implement patient engagement in their study [2, 16]. This training can be time-consuming and may come with a cost depending on the scope of the study. Additionally, previous findings have noted that the type of engagement implemented in a study may pose some challenges. For example, focus groups are a common method to gain the patient perspective, however, patients may lack the confidence to participate in group discussions if they feel they do not have enough to contribute or are concerned with their lack of scientific knowledge. [2, 10]. This form of engagement also may not provide a representative view of all patient perspectives and may present opportunities for conscious or unconscious bias [12, 17]. These challenges have created skepticism surrounding the value of PE efforts and whether the benefits of this approach outweigh the challenges [1, 11, 5]. In addition to these challenges, the lack of guidance surrounding patient engagement or effective practices can add to the difficulty of implementing patient engagement. The role and responsibilities of a researcher and patient in this partnership is often unclear and the power difference between the two groups may pose some hesitancy surrounding the patient’s willingness to share their opinions [8, 18]. A final challenge that has come to light in patient engagement is that of tokenism. In this context, tokenism can be described as the role of the patient being only a symbol to create the false appearance of inclusivity in research [5, 14, 18]. This results in a lack of true engagement and the potential for unequal treatment of patient partners in the research environment [5]. Tokenism also raises the possibility that a research team may devalue patient input [14].

Some researchers have suggested that there is a moral obligation to involve patients in research of topics that pertain to them [4, 19]. For example, Esmail and colleagues proposed that patients have a right to engage as partners in studies that impact them because they are the end-users of the research results [4]. Although the proposed moral obligation of patients to participate in research can cause potential moral tensions, it is valuable to note that the involvement of patients ensures research focuses on the patients’ needs [4, 19].

Previous studies have emphasized the importance of patient engagement in scientific research. Patients bring a new perspective that researchers did not previously have [2]. Involving patients helps to ensure that research topics are based on patient need and that the outcomes are relevant and of value to patients [1, 8]. Relevant findings can be more applicable for use in healthcare practices and are therefore imperative to improve health outcomes [19]. Without patient engagement, studies will likely continue to be driven by the researcher and commercial interests with a lack of regard for patient needs [20]. A literature analysis published by Jun and colleagues in 2018 established that approximately one out of five papers collected between 2010-2013 addressed a priority research topic identified by patients, caregivers, and clinicians in a previously completed research priority setting task. This implies that most of the research within that time frame was not meeting the needs of patients emphasizing the need for greater engagement of patients in research [20].

### 1.5 Knowledge Gap

Actively involving patients in research is a relatively new approach and has been described as a major evolution in healthcare research [3]. In the past, many studies did not consider the patient experience in the development of research questions. Studies were mainly based on what the researcher deemed relevant and important in the context of the specific health condition [21]. Traditionally, clinical trials have not prioritized or reflected upon patient needs, which has led to a disconnect between the research and the issues that are meaningful to patients [5]. Acknowledging the importance of patient engagement in research is a growing movement, alongside the shift toward simpler clinical trials that emphasize enhanced patient care [5].

Although there have been great advances and the approach of patient engagement has been gaining traction in scientific research, there is no gold standard for how to implement the most effective patient engagement. Current literature has noted that more evidence is needed to identify best practices for effectively engaging patients [3, 8, 21]. There is also limited evidence surrounding the impact of engagement on research outcomes and healthcare decisions [3, 8, 14]. Further research investigating the benefits, challenges, importance, and implications of PE from the perspectives of both patients and researchers would enhance the current knowledge base surrounding effective engagement, alluding to the aims and objectives of this current study.

### 1.6 Research Objectives

The objective of this study is to examine the perceived benefits, challenges, importance, and implications of patient engagement efforts from the perspectives of sarcoma patients and the research team. It is hypothesized that patient engagement in research is correlated with increased knowledge of sarcoma research and a greater sense of community for patients. As well, it is predicted that engagement will provide insight to researchers on relevant topics and effective engagement activities. Challenges may include patients’ potential struggle to understand the research study, and the increased time needed for researchers to allocate to engagement efforts. This study uses a mixed model research design comprised of qualitative data collected from a focus group with patients, and a quantitative survey completed by the research team to gain insight on the value and effectiveness of current patient engagement efforts. Overall, the area of research pertaining to patient engagement is of importance because continuous patient engagement can aid in the success of clinical trials and assist in the improvement of the patient experience. Further outlining the value of patient engagement and how the benefits outweigh the potential challenges is an essential step to improving guidelines surrounding the implementation of patient engagement.

## 2.0 Methods

### 2.1 Focus Groups with Patient Advisors

Focus groups have been used as a method of qualitative data collection for many years in the field of health research [22]. This method commonly entails a group discussion on a specific research topic in a safe and natural environment carried out by a moderator or facilitator [22, 23]. Focus groups are a cost-effective way to gain insight into personal opinions, perceptions, attitudes, and experiences from relevant groups of people on the research topic [22]. This approach facilitates the exchange of ideas and opinions which helps to collect individual, group, and interactional data. This data is commonly used to guide the improvement of services in the healthcare system for service users as well [22].

The sarcoma patients involved in this study are patient advisors in a larger Patient-Centered Research (PCR) Advisory Group which assists with the design, implementation, and dissemination of the Surveillance AFter Extremity Tumor surgerY (SAFETY) randomized controlled trial. Purposeful sampling was utilized as the chosen patient advisors had relevant and valuable experiences to share. Patient advisors were initially approached to participate in a focus group via email. Two separate 90-minute virtual focus group discussions were conducted over Zoom due to varying availability. These discussions aimed to collect qualitative data on the benefits, challenges, importance, and implications of patient engagement from the perspective of sarcoma patients. Three participants from the SAFETY PCR Advisory Group attended the first focus group in November of 2021, and 2 participants attended the second focus group in March of 2022. Four members of the SAFETY Trial research team attended both focus groups. An experienced focus group facilitator was also present to lead both discussions. Prior to the start of each discussion, the facilitator reviewed a Participant Information and Consent Form. The same semi-structured focus group guide was used to provide partial guidance for the interactive discussions while also allowing for other topics the participants wished to bring forward (Appendix A). Members of the research team took notes during the discussions on key points, non-verbal behaviour, and the overall group dynamic. The facilitator demonstrated active listening techniques and assisted with the flow of the discussions, ensuring that each participant had an equal opportunity to provide their response to each question. Each focus group session was recorded and later anonymized and transcribed by a transcriptionist.

Discussion items included opinions regarding the SAFETY study such as initial thoughts, clarity of the discussed engagement plan, recommendations for the description of roles in the engagement plan, recount of the patients’ experience as a patient advisor thus far, the effectiveness of the implementation of the engagement plan, strengths and challenges of the engagement plan, and how patient engagement has impacted the SAFETY trial. More general questions included what participants think makes a partnership between a patient and researcher meaningful, the importance of patient engagement in research, feelings associated with being a patient advisor, how patients benefit from being engaged in research, challenges to implementing patient engagement and how these challenges influence patient participation, and other factors that the participants think would help facilitate patient engagement.

### 2.2 Research Team Survey

A survey created on Google forms was used to collect quantitative data on the benefits, challenges, implications, and importance of patient engagement from the perspective of researchers (Appendix B). The survey was completed by four members of the SAFETY trial research team; the principal investigator, research manager, research coordinator, and research assistant.

The survey topics and questions were developed following an extensive review of the literature surrounding patient engagement. Past work outlining the benefits, challenges, implications, and importance of patient engagement assisted in the development of survey answer options to probe the research team’s perspective on current knowledge in this field.

Additionally, a review of previous research allowed for the identification of knowledge gaps in which the developed survey questions aim to address. In developing the survey questions, wording was chosen based on terms previously used in the literature surrounding patient engagement to ensure clarity and cohesiveness with preceding themes. Survey questions and answer options were specific and detailed with the objective of minimizing misinterpretation.

Some survey questions utilized a Likert scale to measure attitudes, opinions, and agreement with statements and aspects pertaining to patient engagement. The Likert scale is one of the most popular response scales used in medical, educational, and psychometric surveys for data collection to understand the attitudes of individuals [24, 25]. A midpoint was included in this study’s survey as research has found that when respondents are equipped with a significant amount of knowledge on the given topic, a midpoint allows participants to express neutrality or an indifferent opinion on the topic [24]. Utilization of Likert-type questions also helped developed a uniform sentence structure of the survey questions.

Survey items included multiple choice questions on the teams’ current level of knowledge and experience surrounding patient engagement, assigning a score to the level of importance of benefits and implications of patient engagement, scoring the difficulty of challenges to implementing patient engagement, and scoring the level of importance of aspects of patient engagement based on what the researchers think would be important to patients. The survey also included how strongly the research team agreed with statements surrounding the improvement of patient engagement and statements surrounding the impact of aspects of patient engagement on the SAFETY Trial. Open-ended written questions included how the researchers think patient engagement will benefit the SAFETY Trial study and any additional comments.

Following initial development, the survey was piloted, and feedback was provided by a clinical professor and research personnel in the Centre for Evidence-Based Orthopaedics at McMaster University. The purpose of piloting the survey prior to it’s distribution was to assess for face validity, to establish if the survey content is suitable for the objective of the survey, and content validity, to ensure the survey measures what it intends to measure. Following this feedback, necessary changes and adaptations were made to the survey prior to collection of survey responses.

### 2.3 Qualitative Data Analysis

Qualitative description (QD) is a common qualitative method that aims to describe a participant’s experiences and perception of a specific research topic [26]. This approach is commonly used to analyze data from semi-structured interviews and focus groups. Data analysis using this method is comprised of a description of a participant’s experience and stays very close to the data by drawing from language used by participants [26]. The focus group transcripts for this study were analyzed using qualitative content analysis. Qualitative content analysis is a commonly used qualitative research technique and aims to interpret meaning found in the context of text data [27]. This conventional analysis approach derives codes and coding categories directly from the text data [27]. Deriving themes directly from the transcript through inductive coding ensures that information is obtained directly from the study participants without preconceived perspectives or potentially biased themes.

The focus group facilitator, research coordinator, and a research assistant completed independent open coding of each independent transcript following the focus group discussion. Then, the research team met to come to a consensus and develop an initial list of codes to apply to the whole transcript for each focus group. This code list evolved as the transcripts were reviewed numerous times. Codes were then organized into meaningful and relevant categories based on the review of each individual’s findings throughout the coding process. This approach of peer debriefing ensures a greater credibility of the data and emerging themes [27]. The number of occurrences of each theme were counted to outline the most common themes.

Following completion of both focus group discussions and related coding, a comprehensive list of codes was created to allow for the extensive analysis of trends in both focus group discussions. Trends from the focus group discussions were used to further analyze the patient perspective on patient engagement.

### 2.4 Quantitative Data Analysis

Survey responses were compiled and organized into an Excel spreadsheet for further analysis. Survey questions involving assigning scores of importance or difficulty were outlined on a scale of one to five, with one being least important or difficult, and five being most important or difficult. Average scores were calculated for each question that involved scorings of importance and difficulty. From these average scores, bar graphs were created to visualize the trends seen in the data. Pie charts were developed to visualize the responses to questions involving how strongly researchers agreed with the presented statements.

## 3.0 Results

### 3.1 Focus Group – Qualitative Findings

Following qualitative coding, the focus group discussions with sarcoma patients revealed many relevant themes related to patient engagement from the patient perspective. Table 1 outlines the most common themes with accompanying quotes that demonstrate these themes. The most common themes with 7 occurrences each were storytelling and communication followed by contribution, helping others, and value with 6 occurrences each. Themes with 5 occurrences included community, experience, involvement, patient perspective, and reassurance. Other themes related to why each participant wished to be a patient advisor centered around the desire to add to research in the field of sarcoma, connect with other patients, and support future patients.

**Table 1.**
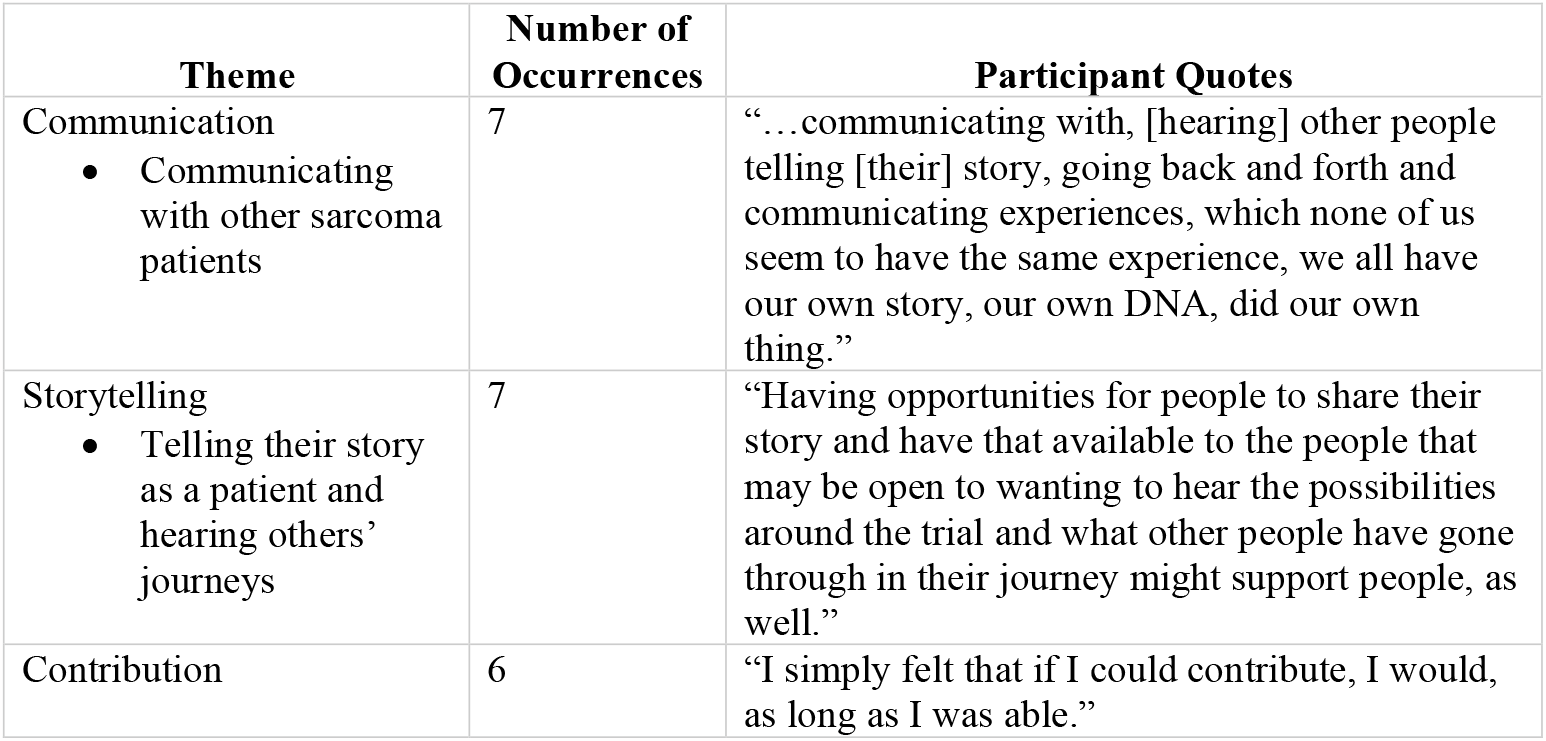
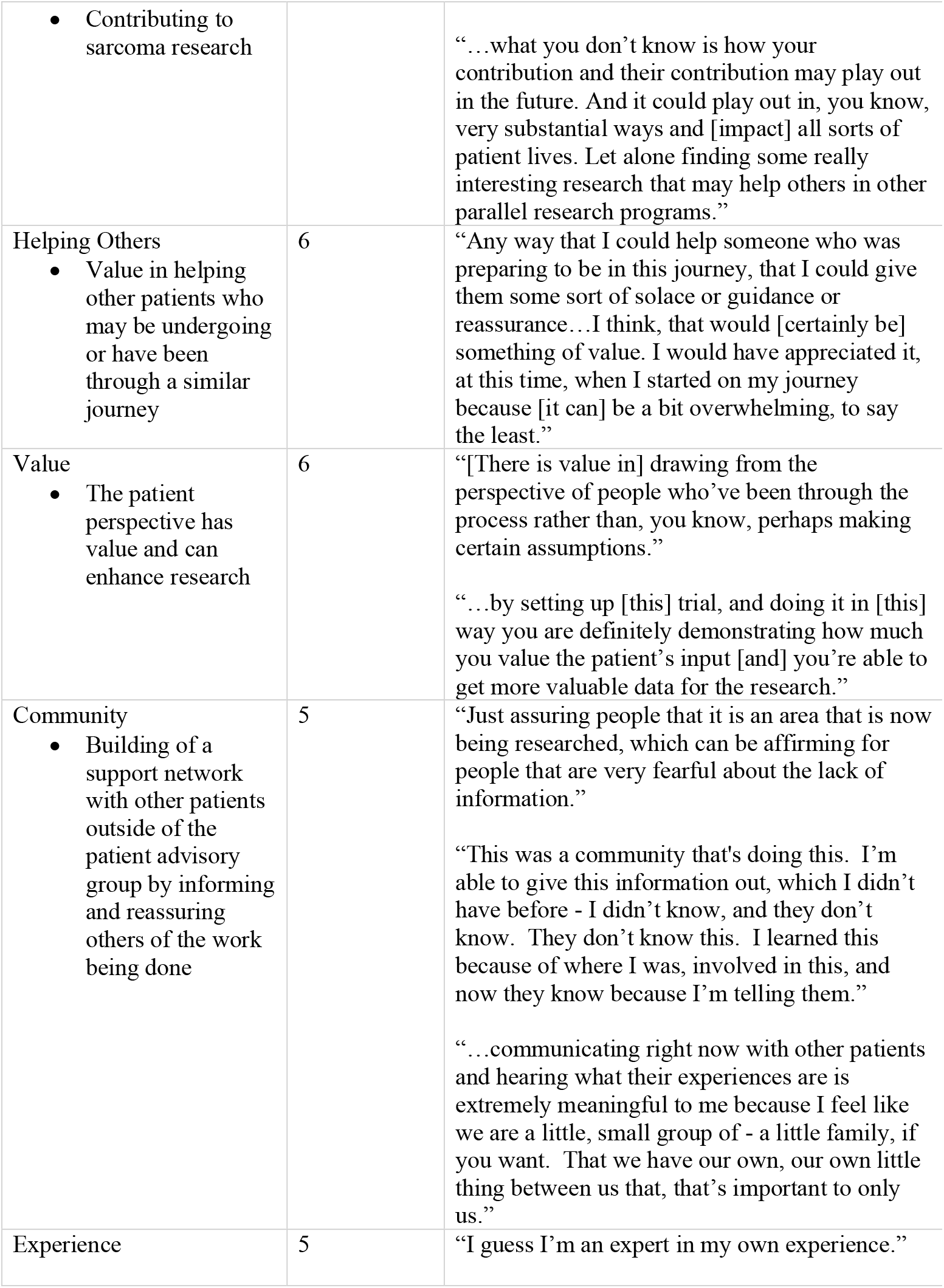
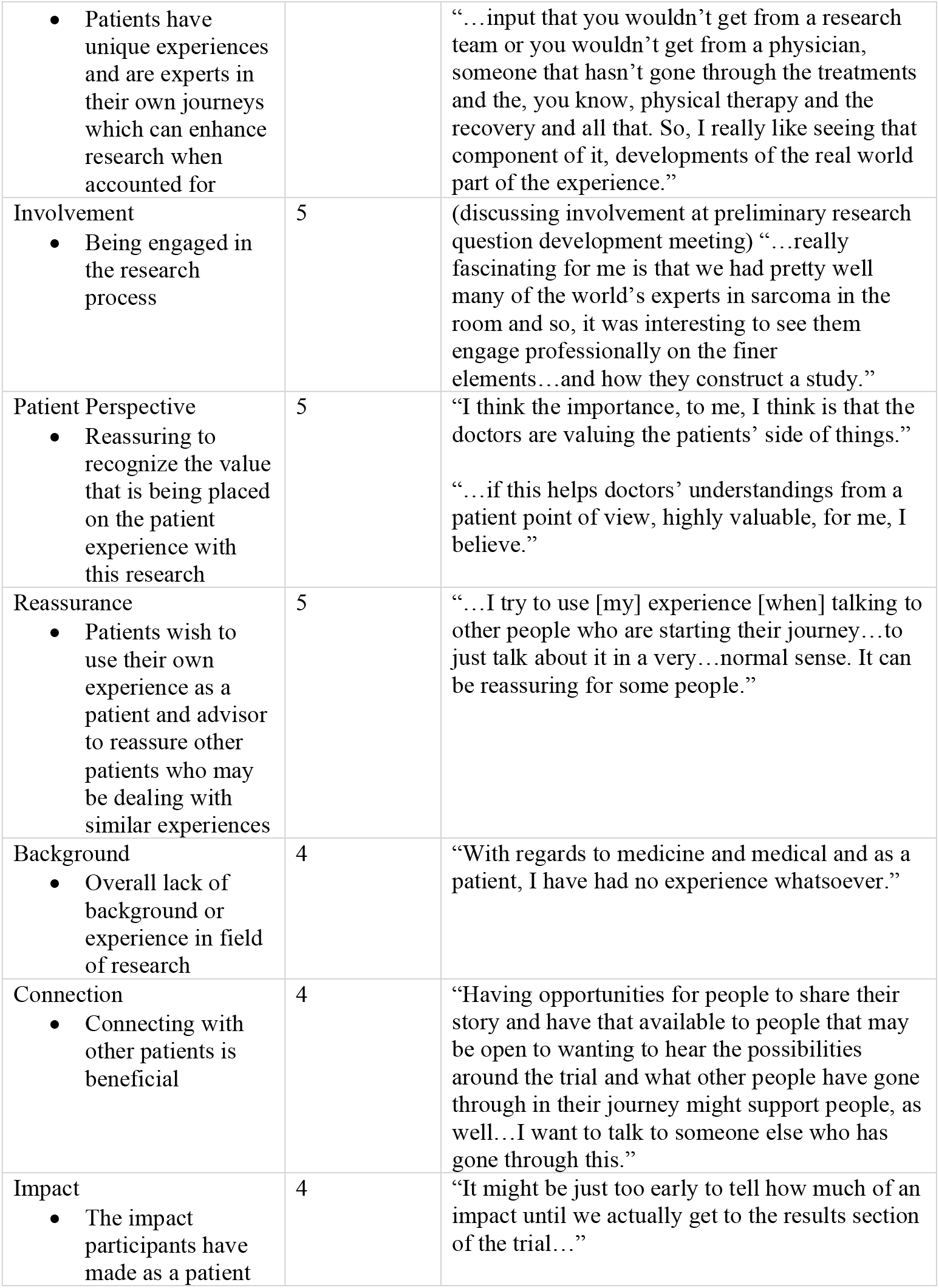
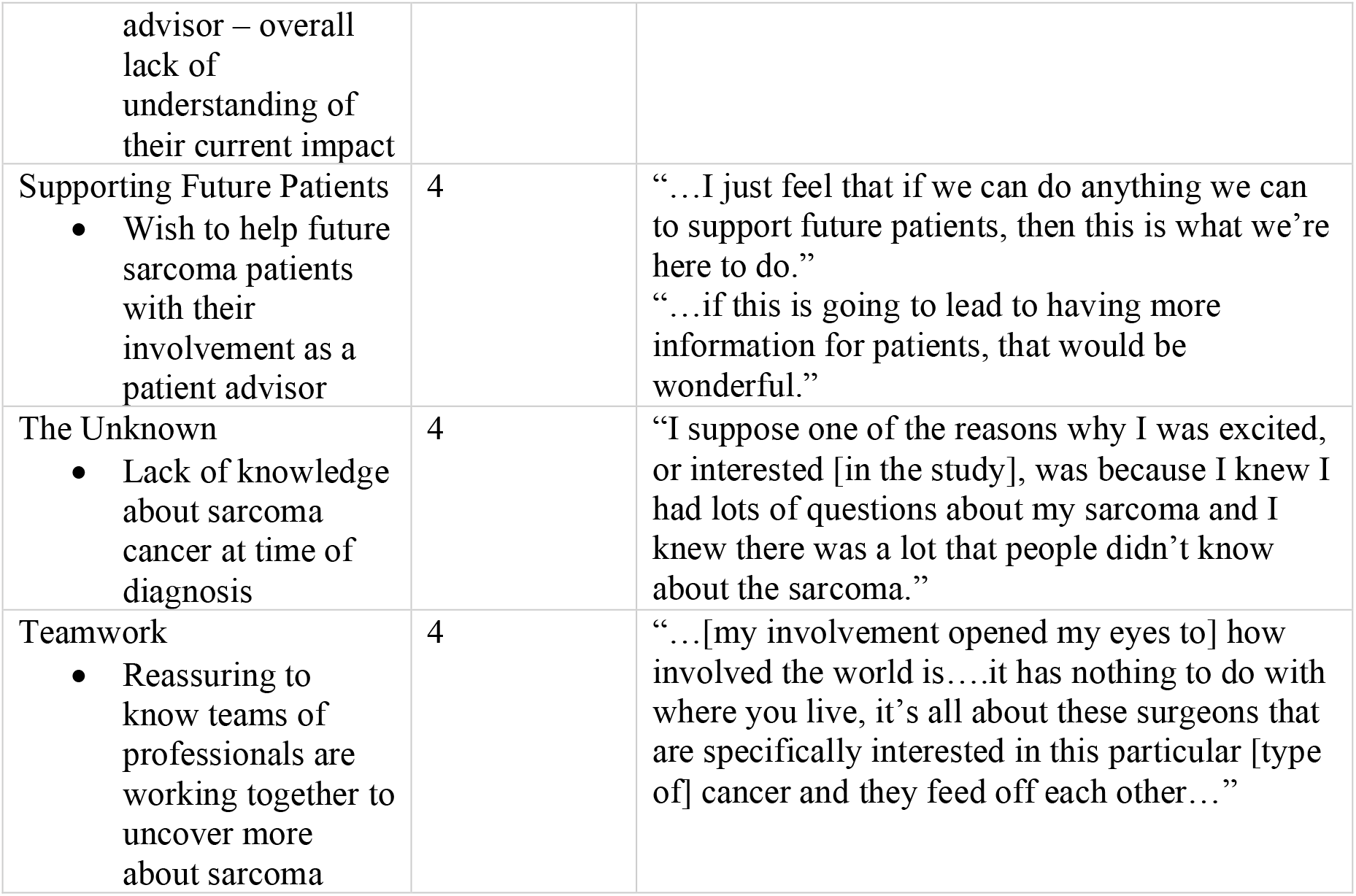
Most commonly occurring themes from the focus group discussions with patient advisors. Following qualitative coding, themes were pulled from the data and the number of occurrences of each theme was counted. The left column outlines the title of the code and a brief note on what the theme entailed based on language used during the focus group discussions. Participant quotes were pulled directly from the transcripts.

Many themes outlined the benefits of patient engagement. These themes included gaining a sense of community by being involved as a patient advisor. Communicating with other patients who have had similar, yet unique journeys was also noted as a cathartic process. Being a patient advisor was referenced as the opportunity to join a supportive family and community with added reassurance that work surrounding this rare type of cancer is being done.

The major challenge to patient engagement mentioned by participants in the focus group discussion was a lack of experience in the science or research field. Some aspects of the current SAFETY Trial patient engagement plan raised confusion for the focus group participants who were introduced to the 10-step framework during the focus group. One participant mentioned “I think having input into the information that is provided to patients and ensuring that it has the common language – [not] eliminating but sort of a better understanding around the medical jargon, potentially.”. Although the plan was geared towards a general audience, the participants were able to identify areas that raised questions due to use of jargon. Additionally, a participant noted a theme of foreign concepts; “I need to really roll it around in my mind sometimes because they’re foreign concepts for me to… to engage in.” This indicates a further challenge due to a lack of background knowledge. Lastly, a theme that arose was a lack of understanding surrounding their impact as a patient advisor in the larger clinical trial thus far. One participant noted that “I’ve come to appreciate that there are many moving parts when it comes to a trial that’s as large and international with many partners…I’m not sure if [the patient advisory group] [or I’ve] made an impact.” Similarly, another advisor mentioned that “It might just be too early to tell how much of an impact until we actually get to the results section of the trial…”

In terms of the importance of patient engagement, many participants noted that at the time of their diagnosis, there was a lack of support for individuals with this form of cancer and a lack of information surrounding their diagnosis. Participants reported the appreciation and reassurance they feel that work was being done in this area after witnessing professionals working together on the SAFETY Trial. The participants discussed the much-needed value professionals are now placing on the patient experience and perspective as a major theme of importance. Broad implications that were noted included the improvements that could be made to patient care with use of the patient perspective, as well as the support of future patients with the findings of the SAFETY Trial.

### 3.2 Research Team Survey – Quantitative Findings

Survey results showed that the average level of current knowledge about patient engagement amongst members of the research team is 3.25 out of 5, or 65%. As shown in Figure 1, it was found that the research team believes that insight into the improvement of patient care practices is the most important benefit of patient engagement with all survey respondents assigning the highest score of importance for this benefit. The benefits of gaining the patient’s perspective to target research towards patient needs and assistance with knowledge dissemination had an average importance of 4.75. Finally, an improvement in enrolment and decrease in attrition scored an average importance of 4.5, and assistance with research question development and study design scored an average importance of 3.75.

**Figure 1.**
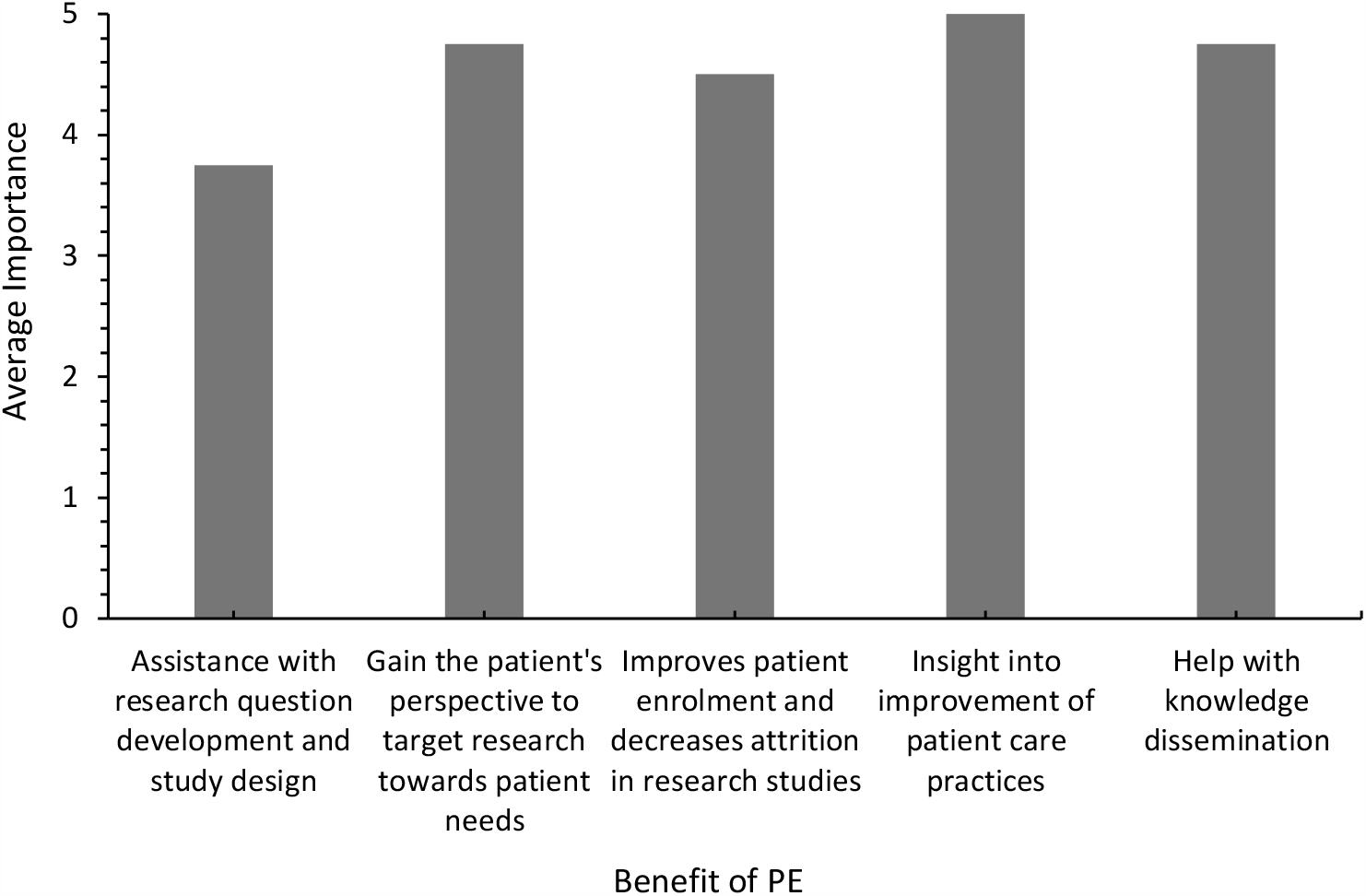
Bar graph displaying responses regarding the importance of various benefits of patient engagement. Options for this survey question ranged from one to five with one being not important, three being somewhat important, and five being very important. The average score for each benefit of PE was calculated to obtain an average level of importance. As shown in the figure, the most important benefit of PE to researchers is gaining insight into the improvement of patient care practices, as displayed by an average importance of 5. The next most important benefits of PE were gaining the patient perspective to target research towards patients’ needs and help with knowledge dissemination, with both having an average importance of 4.75. (n=4).

As seen in Figure 2, the most difficult challenge regarding the implementation of patient engagement was the lack of guidance for PE efforts with an average difficulty score of 4.5. Difficulty selecting patient partners and limited time to develop relationships with patient partners were scored as the next most difficult challenges, with average difficulty scores of 3.75 and 3.5 respectively. An average difficulty score of 3.25 was found for the need to educate patients and communication challenges. Lastly, the least difficult challenge for an average score of 2.75 was added expenses that accompany patient engagement efforts.

**Figure 2.**
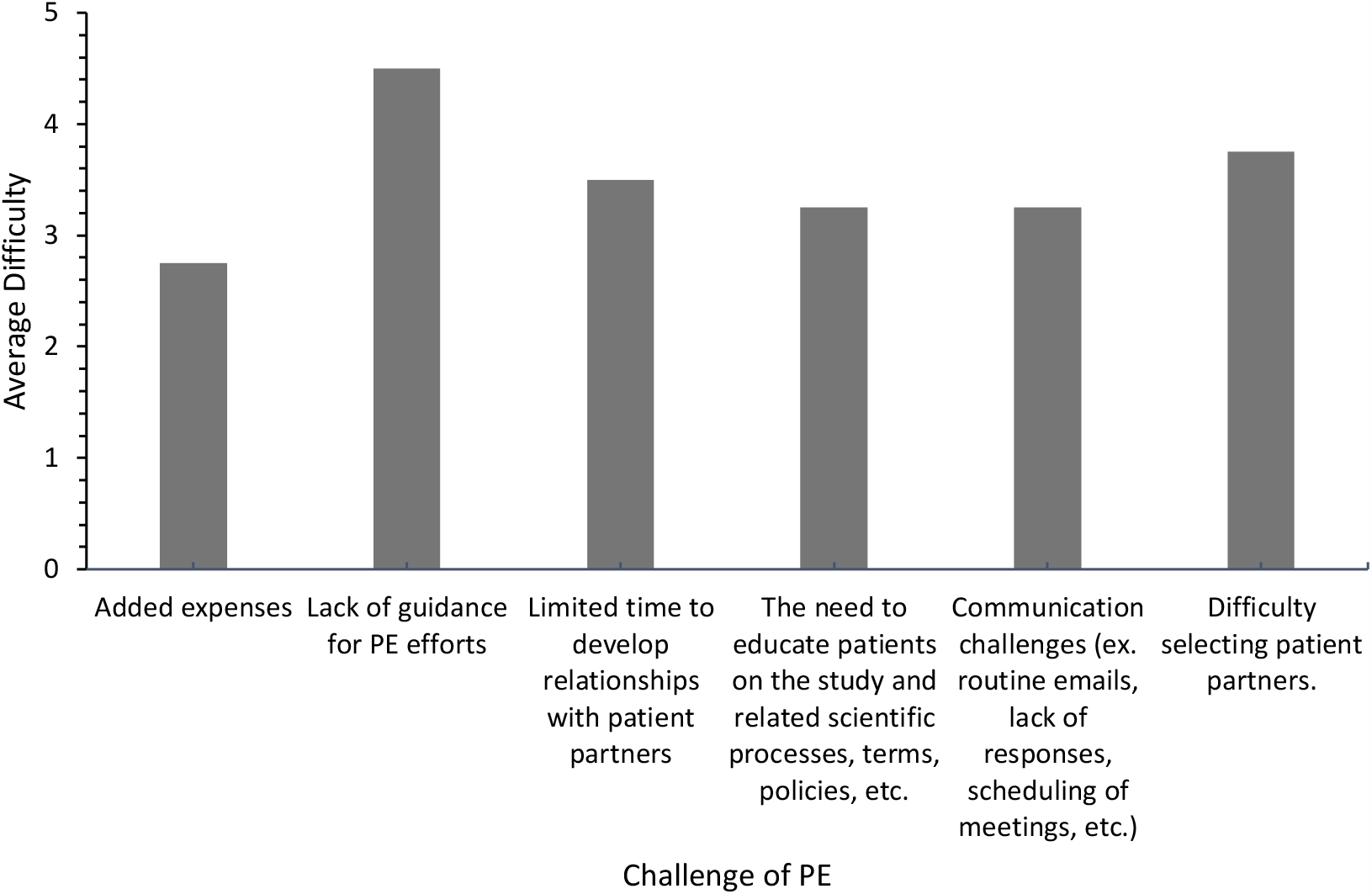
Bar graph displaying responses regarding the level of difficulty of various challenges of patient engagement. Options for this survey question ranged from one to five with one being not difficult, three being somewhat difficult, and five being very difficult. As shown in the figure, lack of guidance for PE efforts was ranked as the most difficult with an average ranking of 4.5, and added expenses was ranked as the least difficult challenge with an average ranking of 2.75. (n=4).

When assigning scores of importance to implications of patient engagement, all implications received very similar average scores as shown in Figure 3. The implication with the highest average score of 4.75 was the use of the patient experience to facilitate the improvement of patient care practices. All other implications such as targeting study outcomes to topics that are relevant to patients, increasing the effectiveness of the translation of research findings, and patient education and empowerment received the same average importance score of 4.5.

**Figure 3.**
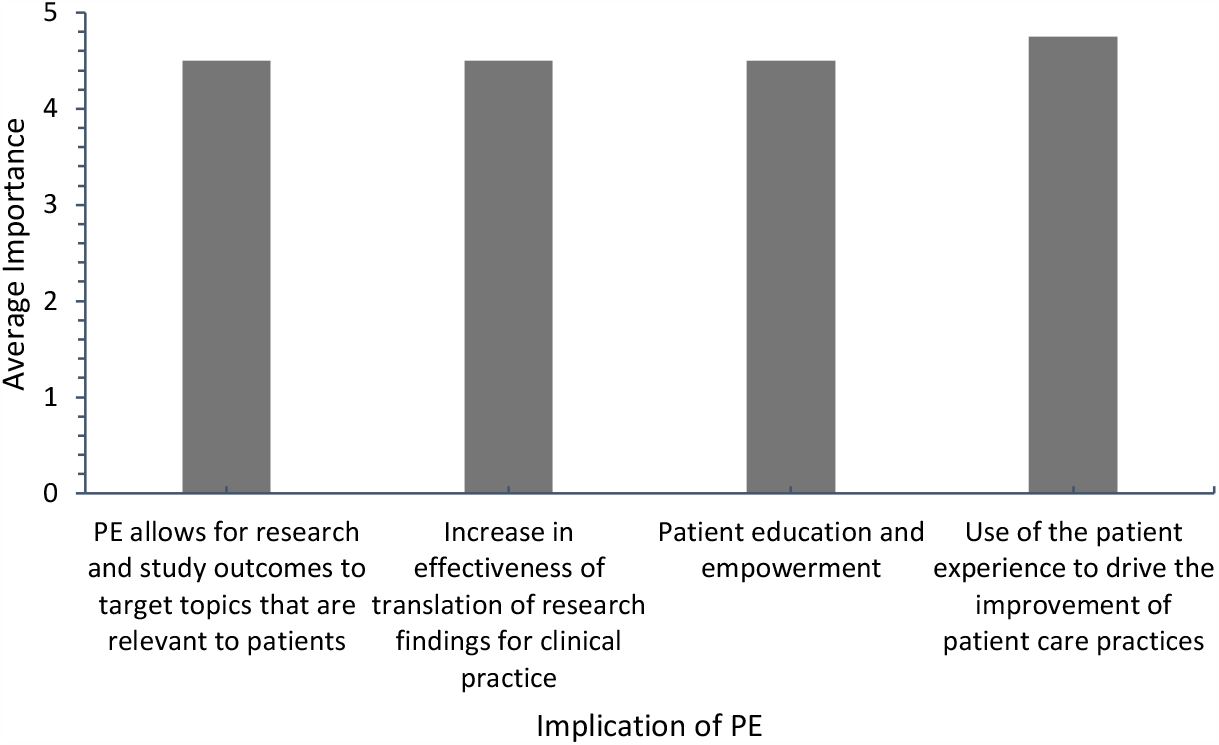
Bar graph displaying responses regarding the level of importance of various implications of patient engagement. Options for this survey question ranged from one to five with one being not important, three being somewhat important, and five being very important. The implication with the highest average importance ranking of 4.75 was the use of the patient experience to facilitate the improvement of patient care practices. All other implications had an average importance ranking of 4.5. (n=4).

When asked to assign a level of importance for aspects of patient engagement based on what the research team thinks is most valuable to patients, three out of the four statements received an average importance score of 4.5 while the remaining statement received an average score of 3.75. The options with an average importance score of 4.5 were gaining a sense of community, helping future patients, and feeling empowered by being involved in research. The aspect receiving an average score of 3.75 was increased knowledge about a disease, treatment, or the research process.

Figure 4 displays the responses to the Likert type survey question involving how strongly the respondent agrees with statements surrounding patient engagement. All members of the research team strongly agreed that there needs to be better guidelines for effective patient engagement practices (Figure 4A). With 3 respondents answering strongly agree and 1 answering agree, results showed that the next most agreed option for improvement was methods to evaluate engagement and the impact of engagement (Figure 4E). With some variability in agreement, the next most agreed upon option was the need for reference material that outlines the responsibilities of both the patient and researcher (Figure 4B). Available funding as a means of improving patient engagement efforts followed with 1 response of strong agreement, 2 responses of agreement, and 1 response of neutrality (Figure 4D). The statement with the most neutrality was that there should be more autonomy given to the patient and researcher in terms of roles and responsibilities (Figure 4C). Lastly, the statement with 1 response of strongly disagree and 3 responses of disagree was the need for patients to have prior research experience or relevant knowledge on the topic to be a patient partner (Figure 4F).

**Figure 4.**
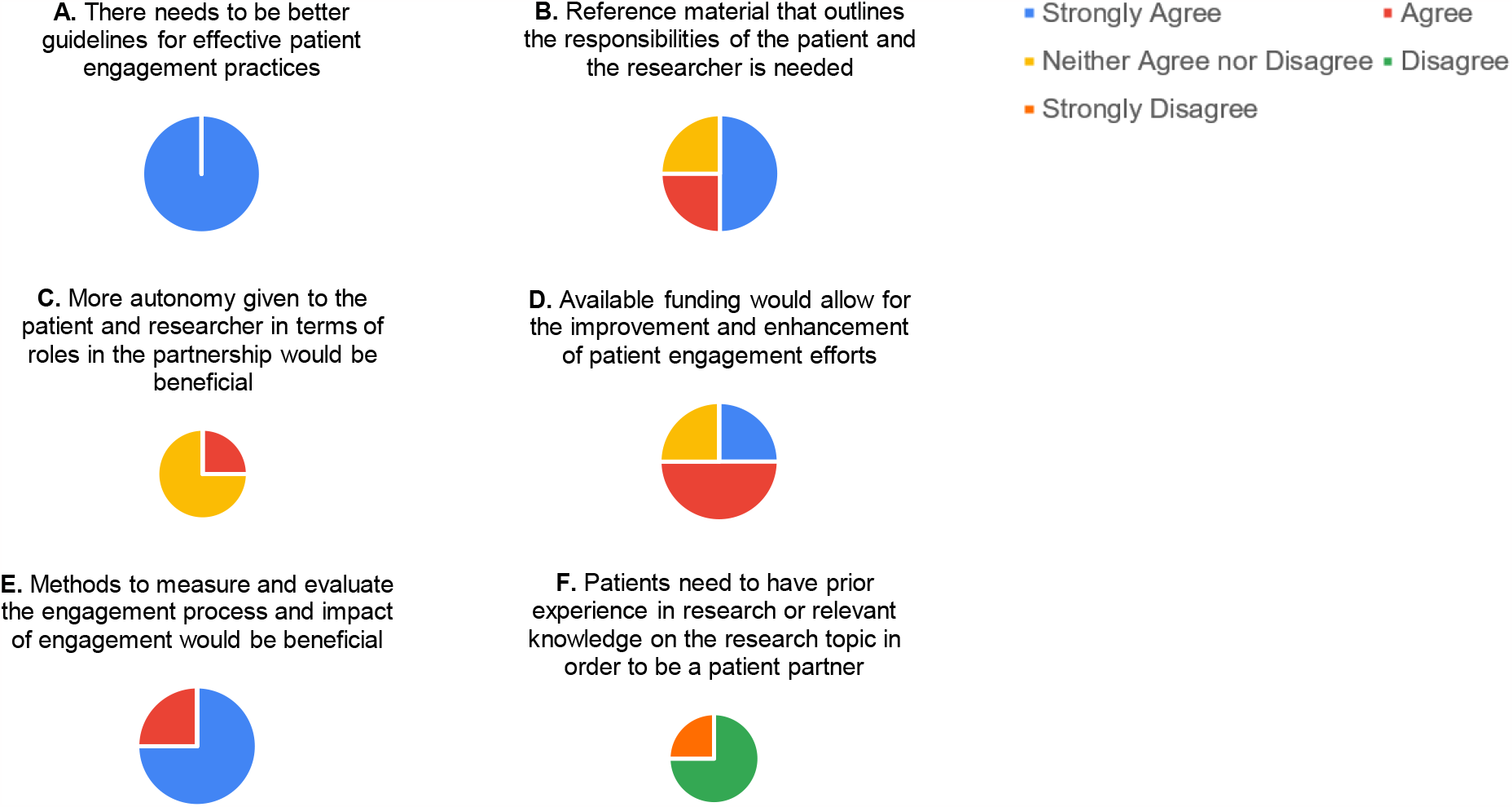
Pie charts displaying the strength of agreement with various statements surrounding the improvement of patient engagement. This survey question utilized a 5-item Likert scale with options being strongly agree, agree, neither agree nor disagree, disagree, and strongly disagree. The statements with the strongest agreement amongst the research team involves the need for better guidelines for patient engagement practices and methods to evaluate the impact of engagement (A,E). The most neutrality was seen surrounding more autonomy given to the patient and researcher (C). A mix of agreement and neutrality were seen for the need for reference material that outlines the responsibilities of the patient and researcher and the improvement of patient engagement with available funding (B, D). The most disagreement was seen regarding the need for patients to have prior research experience (F). (n=4).

Survey responses from the research team showed that their average level of confidence surrounding the effectiveness of current SAFETY Trial patient engagement efforts was 3.75 out of a possible score of 5. To examine this further, the research team was asked to indicate their strength of agreement with statements surrounding the impact of aspects of PE on the SAFETY Trial as seen in Figure 5. Two of the six statements showed the strongest agreement with 3 responses of strongly agree and 1 response of agree. These options were that PE has allowed for the selection of a patient relevant research topic (Figure 5A) and that PE in the SAFETY trial has changed the way members of the research team view the outcomes of the clinical research study (Figure 5B). With 50% of responses being strongly agree and the other 50% being agree, the next most agreed with statement was that patient engagement has enhanced and made the SAFETY trial a more robust study (Figure 5C). Survey results showed that for the impact of improved dissemination of SAFETY Trial findings with patient engagement, 3 out of 4 responses were agree and 1 response was strongly agree (Figure 5E). In terms of disagreement, 50% neutrality, 25% disagreement, and 25% strong disagreement for the statement that PE efforts have added more difficulty to the SAFETY trial was seen (Figure 5D). Strong disagreement from all respondents was noted for the statement surrounding the SAFETY Trial being a more successful study without the involvement of patient partners (Figure 5F).

**Figure 5.**
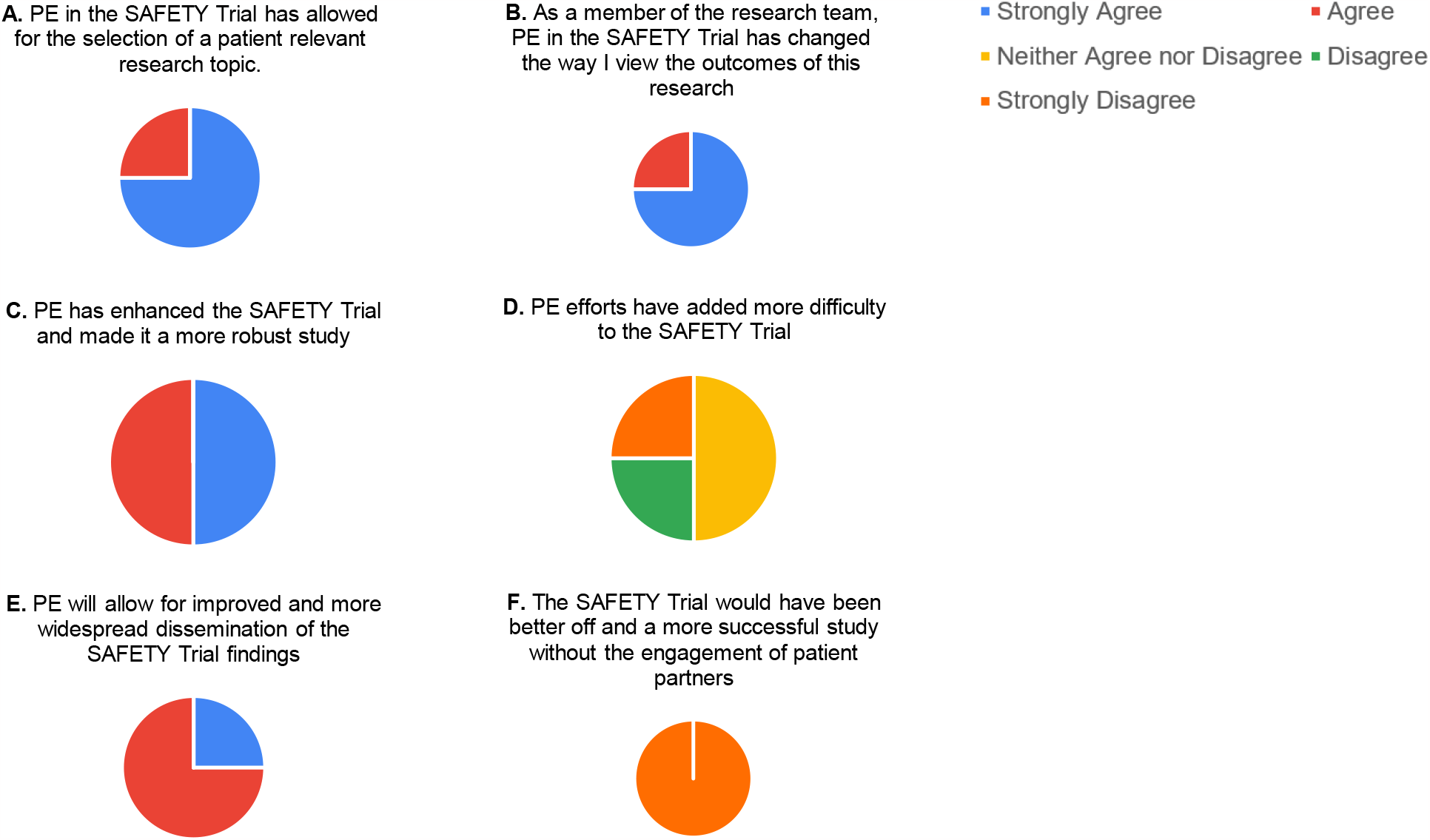
Pie charts displaying the strength of agreement with various statements surrounding the impact of aspects of patient engagement on the larger SAFETY Trial. This survey question utilized a 5-item Likert scale with options being strongly agree, agree, neither agree nor disagree, disagree, and strongly disagree. A mix of strong agreement and agreement were seen for the majority of statements in this survey question (A, B, C, E). Neutrality and agreement were seen for the addition of difficulty to the SAFETY trial with patient engagement (D). The strongest disagreement was seen for the statement positing that the SAFETY Trial would be more successful without patient engagement (F). (n=4).

The survey concluded with an open-ended long answer question inquiring how the researchers think patient engagement has or will benefit the SAFETY Trial study. Themes in the responses to this question are noted along with select quotes in Table 2 below. Overall, there was a consensus that patient engagement assists with identifying topics that are important to patients. Gaining the patient perspective also provides insight into the type and frequency of follow-up patients would prefer. One survey respondent noted that although engaging patients has come with a learning curve, the involvement of patients will enhance the study’s relevance and positively impact clinical practices. In the final survey section for additional comments, respondents noted the lack of guidance for patient engagement activities which adds to the difficulty of implementing patient engagement. It is also important to consider the demographic that volunteers to participate in patient engagement and the generalizability of findings from this patient demographic.

**Table 2.** Open-ended survey question responses. This table outlines common themes surrounding patient engagement found in the open-ended survey questions following qualitative coding. (n=4).

| Theme | Quote |
| --- | --- |
| Identifying what is important to patients | "Hearing patient perspectives allows us to identify what is important to them, and what they want out of their care." |
| Enhancing clinical practices | "PE will help us understand how patients feel about the different treatment arms. This will guide any changes that clinicians will make in their practice when determining follow-up frequency/type." |
| Assistance with study design | "Patient engagement has informed the study question and trial design and continues to inform the study execution." |
| Knowledge translation | "I think it will be of particular benefit in the knowledge dissemination phase to ensure the results are accessible to both healthcare workers and patients." |

## 4.0 Discussion

### 4.1 Benefits

This study reinforces and adds to the current literature surrounding the benefits of patient engagement from both the patient and researcher’s perspective. Previous research has concluded that a major benefit of PE is gaining a perspective from an individual with lived experience to assist in the design of a research study [1]. This benefit was seen in the ‘Patient Perspective’ and ‘Involvement’ themes in the focus group discussion (Table 1). From a researcher’s perspective, this benefit was ranked as the second most important benefit of PE (Figure 1). The research team also noted in their written survey responses that hearing the patient perspective allows for the identification of topics that are important to patients (Table 2). This coincides with findings from Manafo and colleagues surrounding the improvement of the applicability of research outcomes with the inclusion of patient perspectives [3]. Manafo and colleagues also noted that PE helps to increase study enrolment and decrease attrition as patients are being actively engaged and feel more inclined to participate when the study outcomes are of value to them [3]. The average level of importance for this benefit of PE from the perspective of the research team was 4.5/5, indicating that this aspect is a large contributor to the value of implementing patient engagement.

A prominent theme that arose from the patient perspective was that of ‘Storytelling’, ‘Communication’, ‘Community’, and ‘Connection’ (Table 1). These themes mainly entailed the opportunity patients had to share their story, listen to others’ stories, and overall build a support network with individuals who have followed a similar journey. These findings emphasize the need for enhanced support of patients as they navigate their diagnosis, treatment, and follow-up and the essential component that PE adds to the objective of improving the patient experience. Previous work by various researchers noted the empowerment patients gain by being involved in research [1, 3, 4, 15]. This empowerment can be seen as a byproduct of the previously outlined themes from the focus group discussion as focus group participants agreed that being part of the patient advisory group felt cathartic as it provided the chance to speak to others with a certain level of understanding surrounding the patient journey with sarcoma. Additionally, ‘Supporting Future Patients’ can be tied to feelings of empowerment and was noted as a major driving factor for patients to participate in the current trial as a patient advisor.

Similarities were observed between the perspectives of patients and the research team regarding some benefits. Both patients and the research team value the benefit of acknowledging and communicating the patient perspective to advance research and patient care. As expected, from the patient perspective, most benefits of PE surrounded communication, sharing stories, and supporting future patients. This aligned with what the research team believed to be important to patients, indicating a relatively cohesive vision of the objectives of patient engagement for the patient advisors and research professionals participating in this study. In contrast, patients in the focus group noted one benefit to their involvement as a patient advisor being uncovering some of the unknowns they were faced with during their diagnosis. This can be an insightful theme that signals to researchers and healthcare professionals that patients diagnosed with rare cancers need to be equipped with more knowledge surrounding their condition which can also promote enhanced mental wellbeing throughout the patient’s journey.

Overall, this work solidifies the broad benefits of patient engagement from the perspective of relevant groups and coincides with the hypothesis of this study. This further emphasizes the value of patient engagement and provides evidence for the need for PE despite the potential challenges discussed below.

### 4.2 Challenges

A major challenge noted in prior literature is the complexity that comes with incorporating aspects of patient engagement in clinical research [1, 2, 5]. However, survey responses in this current study showed neutrality and disagreement for this challenge (Figure 5). As well, contrary to previous findings, added expenses that may come with patient engagement was not noted as a significant challenge by the research team (Figure 2). On the other hand, training and guidance for both patients and researchers is essential for successful PE as shown in previous research, however, as noted by the research team’s survey responses, there is a current lack of guidance for effective implementation of PE efforts. This challenge was noted as adding the most difficulty to PE implementation (Figure 2). Additionally, there was a consensus among focus group participants that they were unaware of their direct impact as a patient advisor, further highlighting a potential lack of structure of the SAFETY Trial PE plan. This lack of guidance also strengthens the findings of previous literature surrounding the unclear roles of a researcher and patient in a professional relationship involving engagement [8]. Although this is a significant challenge, added research pertaining to what makes PE most successful can help to minimize this difficulty and maximize upon the benefits of PE.

Another challenge noted by the research team was the difficulty of selecting patient partners (Figure 2). Although random sampling would be ideal, this is often difficult with a small sample of patients and the increased inclination some patients may have to participate in comparison to others. Proven in previous literature, all individuals are subject to conscious or unconscious bias which may play a part in studies involving patient engagement and impact the level of impartiality needed to gain accurate results surrounding a given study topic [12]. This challenge emphasizes the need for stricter guidelines regarding the process of choosing patient advisors.

A final challenge discussed in the literature that was relevant in this study is that of tokenism. Tokenism describes the role of the patient as only a symbol to create an appearance of inclusivity in research resulting in a lack of true engagement [5, 14, 18]. This challenge was noted in a written response by a research team member. This respondent stated that “It has been a steep learning curve to become reasonably confident with the involvement of patients in the research continuum, with lots of consideration on how PE can be meaningfully involved (i.e., not tokenistic).” This quote portrays the conscious actions of the research team to avoid tokenism as well as the challenge and learning curve that this imposes. Stronger guidelines surrounding patient engagement can mediate this challenge and help researchers feel more confident in their abilities to effectively engage patients.

Overall, challenges of patient engagement noted by patients differed from those ranked most difficult by researchers. As discussed in the focus group, from the patient perspective challenges arise from their lack of scientific knowledge and the research process. For the research team, the most difficult challenge is the lack of guidance surrounding the implementation of PE efforts. Both challenges stem from a lack of education, either surrounding science as a patient or guidance as a researcher. Along with the other noted challenges, it is essential to consider the overarching cause of these challenges in efforts to mediate these barriers for future patient engagement endeavours.

### 4.3 Importance and Implications

The findings from this study have highlighted the value of patient engagement to both patients and researchers. Although challenges to this approach are noted, it is abundantly clear that the approach enhances research and has implications worth overcoming the difficult aspects. With the benefits and implications that arose during the focus group discussion and that were ranked in the survey, it can be concluded that patient engagement allows for study outcomes to be targeted to patient needs which can assist in the improvement of patient care practices (Table 1, Figure 3). This has been previously found in literature as patients bring a new perspective that researchers do not have which has proven to be helpful for both the patient as findings are of value to them, and the researcher as insight into the patient experience helps to guide patient care [1, 2, 8]. Without patient engagement, it is likely that studies will continue to be driven by researcher interests as topics that are relevant to patients would not get identified. Patient engagement implementation allows for more meaningful research, as mentioned by a research team member in the open-ended survey questions, and helps to further demonstrate the importance of clinical research such as the SAFETY Trial. As noted in Figure 5, the research team agrees that patient engagement has allowed for a more robust clinical study and has changed the way the research outcomes of this work are viewed. The knowledge gained from this study will help in the facilitation of the much-needed improvements to current patient engagement efforts.

### 4.4 Limitations and Next Steps

One limitation of this study is the small sample size of the focus group discussions with patient advisors. The total sample size for the focus group discussions was 5 patient advisors. It is important to be cognisant of the potential lack of generalizability of the findings from the focus group discussion due to this small sample size and relatively similar demographic characteristics of each patient advisor. Individuals of different ethnic backgrounds, academic backgrounds, and socioeconomic statuses may have different views surrounding patient engagement and the general nature of clinical research. The length of time between focus group 1 and focus group 2 may have also impacted advisor’s views and opinions surrounding patient engagement as those that participated in focus group 2 were involved as patient advisors longer than those that participated in focus group 1. To mediate this limitation in future studies involving focus group discussions, one focus group can be conducted instead of two separate discussions to ensure the variable of time is controlled for amongst the participants.

Although each member of the research team conducting the larger clinical trial completed the survey for this study, another potential limitation is the small sample size of this research team. There was a lack of strong statistical testing options available for the ordinal survey data due to this limitation. Next steps to mediate these limitations is future research that gains the perspectives from a larger patient demographic and more researchers involved in other clinical trials with patient engagement components. This would allow for greater generalizability of findings and the opportunity for the use of different quantitative statistical tests to analyze survey data.

Additionally, results from this study can assist the research team in their present patient engagement efforts with the SAFETY Trial. This study highlights that researchers have a lot to learn from patients, and both parties benefit with effective patient engagement strategies. With challenges to patient engagement identified in this study, the research team can begin to find ways to overcome these barriers with the help and guidance of the patient advisory group. For example, with an area of difficulty identified as the lack of scientific knowledge as a patient, the research team can help to mediate this barrier by ensuring all material provided to patients is conveyed in an accessible manner. Further action items include maximizing upon the benefits of patient engagement by continuing to use the patient perspective to improve healthcare practices. Utilizing input from patient advisors can help to identify patient-relevant study topics and care practices to improve future patients’ journeys following their diagnosis.

## 5.0 Conclusion

In conclusion, patient engagement is a valuable approach used in clinical research that aims to involve patients in each step of the research process to enhance the study design and increase the relevancy of the study findings for patients. These present findings coincide with the hypotheses of this study. In past research and much of current research, there is a lack of emphasis placed on the patient’s journey. Gaining insight into the patient experience is crucial to the improvement of patient care practices and ensures that the findings of research can be smoothly translated into clinical practice. This was observed in this study as both patients and the research team conveyed that they strongly valued the use of the patient perspective to help future patients and improve on current practices. Findings from this present survey will assist the SAFETY Trial research team successfully implement patient engagement practices and will allow for a precedent to be set surrounding the investment into patient engagement efforts. Future studies should continue the exploration of patient engagement to further demonstrate the need for this approach and outline best practices. Effective and continuous patient engagement is crucial to the success of clinical research and guidelines that highlight the adoption of a patient-centered approach are needed to ensure maximal benefit from patient engagement efforts.

## Data Availability

All data produced in the present study are available upon reasonable request to the authors

## Appendix

### A. Focus Group Discussion Guide

<u>..\CEO Oncology\Focus Group Material\Focus Group Prep\SAFETY Trial - Patient Engagement Sub-Study - Discussion Guide - V2.0 FINAL 21SEP21 (002).pdf</u>

### B. Survey Distributed to the Research Team

<u>..\CEO Oncology\Survey for Research Team\CEO Musculoskeletal Oncology Patient Engagement Survey.pdf</u>

